# Effect of Wearing Glasses for Prevention of SARS-CoV-2 on Visits to Health Care Providers - Additional Results from a Randomized Controlled Trial

**DOI:** 10.1101/2024.08.29.24311868

**Authors:** Ingeborg Hess Elgersma, Petter Elstrøm, Lars G. Hemkens, Arnfinn Helleve, Oliver Kacelnik, Atle Fretheim

**Author notes:** Corresponding author: Ingeborg Hess Elgersma; Folkehelseinstituttet, Postboks 222 Skøyen, 0213 OSLO, Norway.

## Abstract

We previously published results of a pragmatic randomized trial with 3717 participants in Norway that assessed the effect of wearing glasses on the risk of being infected with SARS-CoV-2 and other respiratory infections. Here we present unpublished findings on pre-specified secondary endpoints relying on routinely collected data from Norwegian health registries: Visits to health care providers for any cause (within 21 days), for respiratory symptoms (day 3-28), and for injuries (within 21 days).

1469 participants (39.5%) visited a health care provider for any cause at least once. There was no statistically significant difference between groups, with 39.9.% of participants wearing glasses versus 39.1% in the control group received primary or specialist care (absolute risk difference 0.76%; 95% CI, -2.4% to 3.9%; relative risk 1.02; 95% CI, 0.94 to 1.10).

Similarly, there was no statistically significant difference in visits due to respiratory symptoms (326 participants; 9.1% versus 8.4%; absolute risk difference 0.71%; 95% CI, -1.1% to 2.5%; relative risk 1.08; 95% CI, 0.88 to 1.33).

There were 3 participants having visited primary or specialist care and having an injury registered in a registry for injuries, the difference between groups not statistically significant. The difference between participants in the proportion of visits for injury related diagnoses was nominally statistically significant (1.3% versus 0.7%; absolute risk difference 0.7%; 95% CI, 0.01% to 1.3%; relative risk 1.94; 95% CI, 1.01 to 3.89).

The study underscores the potential of utilizing registry-based outcomes for randomized evaluations of public health measures for infection control.

## Introduction

This follow-up study report presents additional data collected since our initial publication of a randomized trial that assessed whether wearing glasses reduces the risk of being infected with SARS-CoV-2 and other respiratory viruses. In the initial findings from this trial, wearing glasses in the community was not protective regarding the primary outcome of a reported positive COVID-19 test^1^. However, a secondary outcome, the risk of respiratory infections based on self-reported symptoms was lower in the intervention group (absolute risk difference, -3.3%; 95% CI, -6.3% to -0.3%; relative risk, 0.83; 95% CI, 0.69-1.00). Visits to health care providers for respiratory symptoms, injuries, and all causes were pre-specified in the original trial protocol as secondary outcomes^2^, but this data has so far not been available to us. We have now obtained this data from national registries, namely the Norwegian Registry for Primary Health Care (KPR) and the Norwegian Patient Registry (NPR).

The trial is registered at ClinicalTrials.gov (NCT05217797).

## Methods

For this analysis, we included all participants as described in Fretheim et al^1^. The trial was conducted in Norway February 2 to April 24, 2022. We analyzed the proportion of participants having visited health care providers for any cause (day 1 to 21), for respiratory symptoms (day 3 to 28), and for injuries (day 1 to 21) collected from the primary and specialist health care providers within the public health service (KPR and NPR, respectively).

How to assess the association between wearing glasses and the risk for medical care for injuries was not prespecified in the protocol. From the registries we had access to two distinct sources: The Common Minimum Dataset (FMDS) of injuries in NPR^1^ and consultations in primary and specialist care with an injury related diagnosis. We therefore report two measures for injury risk: (1) The proportion of participants with an injury in the FMDS and with associated consultations in KPR and NPR, and (2) the proportion of participants having visited a health care provider for any injury related diagnoses without a preceding injury reported in the FMDS. Of note, such visits also include treatment of injuries having occurred prior to the inclusion of the participant in the study.

Since having visited a health care provider at least once indicates that the individual has experienced the outcome of interest, we only counted the first visit and calculated risk ratios.

## Results

In total 3717 participants took part in the trial (see CONSORT flow diagram in Supplement 1). Of these, 1469 had at least one contact registered in KPR or NPR (1346 in KPR and 287 in NPR, day 1 to day 28). There was no significant difference in the risk of at least one visit to a health care provider for any cause, 739 of 1852 (40%) participants in the intervention group and 730 of 1865 (39%) participants in the control group (absolute risk difference, 0.76%; 95% CI, -2.4% to 3.9%; relative risk, 1.02; 95% CI, 0.94-1.10) (see Table 1).

**Table 1:** Main findings.

| **Outcome** | **Intervention group, n=1852<sup>1</sup>** | **Control group, n=1865<sup>1</sup>** | **Absolute risk difference<sup>2</sup> (95% CI)<sup>3</sup>** | **Relative risk (95% CI)<sup>3</sup>** |
| --- | --- | --- | --- | --- |
| Any visit to a health care provider (day 1-21) | 739 (40%) | 730 (39%) | 0.76% (-2.4%, 3.9%) | 1.02 (0.94, 1.10) |
| Visits to health care providers for respiratory symptoms (day 3-28)<sup>4</sup> | 169 (9.1%) | 157 (8.4%) | 0.71% (-1.1%, 2.5%) | 1.08 (0.88, 1.33) |
| Visits to health care providers for injuries (day 1-21)<sup>5</sup> | 2 (0.1%) | 1 (<0.1%) | 0.05% (-0.13%, 0.24%) | 2.01 (0.19, 43.3) |
| Visits to health care providers for injury related diagnoses (day 1-21)<sup>6</sup> | 25 (1.3%) | 13 (0.7%) | 0.65% (0.01%, 1.3%) | 1.94 (1.01, 3.89) |

<sup>1</sup>n (%)
<sup>2</sup>Regression least-squares mean difference
<sup>3</sup>CI = Confidence Interval
<sup>4</sup>All visits with diagnoses ICD-10 codes J00-J06, J09-J18 and J20-J22 and ICPC-2 codes A03, A77, F70, F72, F73, H71, R01-R23, R29, R71-R83, R99, R991 and R992
<sup>5</sup>Reported injury to the FMDS and ≥ 1 visits for diagnoses ICD-10 codes chapter XIX and ICPC-2 codes A80, A81, F75-F79, L72-L81, N79-N81 and S17-S19
<sup>6</sup>All visits with diagnoses ICD-10 codes chapter XIX and ICPC-2 codes A80, A81, F75-F79, L72-L81, N79-N81 and S17-S19

Similarly, we observed no statistically significant difference in the risk of at least one visit to a health care provider related to symptoms of respiratory infection. In the intervention group 169 of 1852 participants (9.1%) visited a health care provider, compared to 157 of 1865 (8.4%) in the control group (absolute risk difference, 0.71%; 95% CI, -1.1% to 2.5%; relative risk, 1.08; 95% CI, 0.88-1.33) (see Table 1).

In total 3 participants had an injury reported to the FMDS during the follow-up period and also visited primary or specialist care for their injury, 2 (0.1%) participants in the intervention group and 1 (<0.1%) participant in the control group, the difference is not statistically significant (absolute risk difference, 0.05%; 95% CI, -0.13% to 0.24%; relative risk, 2.01; 95% CI, 0.19-43). In terms of all health care use potentially pertaining to injuries, more such visits were found in the intervention group (absolute risk difference, 0.65%; 95% CI, 0.01% to 1.3%; relative risk, 1.94; 95% CI, 1.01-3.89) (see Table 1).

## Discussion

We found no statistically significant reduction of respiratory infection symptoms that led to a health care provider contact. However, the effect estimate was very uncertain. The results were compatible with a relative risk increase by 33% and a relative risk reduction by 12%, which would also be compatible with the previously reported relative risk reduction of self-reported symptoms by 17% (95% CI 0% to -31%).

There was a slightly higher absolute risk of having visited a health care provider for an injury related diagnoses for the participants wearing glasses. We cannot rule out that this indicates a safety signal for wearing glasses, however there were few cases and the uncertainty around the estimate is high.

A surprise finding, to us, was that almost 9% of the participants had been in contact with the health care services for respiratory infection symptoms (day 3 to day 28). The proportion is similar to the proportion reported having had symptoms of respiratory infections in the end of study survey, although the length of follow-up differs between these two endpoints (day 1-17 versus day 3-28). Overall, these findings underline the feasibility of utilizing registry-based outcomes in randomized evaluations of public health and social measures for infection control.

## Data Availability

The datasets generated and/or analysed during the current study are not publicly available due to data protection reasons but deidentified participant data are available in from the corresponding author on reasonable request.

## Declarations

## Ethics approval and consent to participate

The trial was approved by the Regional Ethics Committee of South-East Norway (reference no. 428685).

## Consent for publication

Not applicable

## Authors’ contribution

AF, PE and IHE conceptualized and designed the study. IHE analyzed the data with supervision from PE. IHE, AF and LGH drafted the manuscript. All authors critically revised the manuscript.

## Acknowledgements

Not applicable

## Footnotes

1 An injury in the FMDS is defined as an acute or sudden impact on the body from physical agents, such as mechanical energy, heat, electricity, chemicals, and radiation, and in an amount or size that exceeds the human organism’s tolerance level and/or a sudden absence of necessary agents such as oxygen or heat (as in drowning and freezing). Reporting of personal injury to the FMDS is mandatory for all somatic hospitals that receive injury cases, as well as for some municipal emergency clinics (Bergen and Trondheim)

