## Supplementary figures and images for "Effect of Wearing Glasses for Prevention of SARS-CoV-2 on Visits to Health Care Providers - Additional Results from a Randomized Controlled Trial"

### Supplement 1

**Supplement 2**

*Figure e1: CONSORT flow diagram of participants*

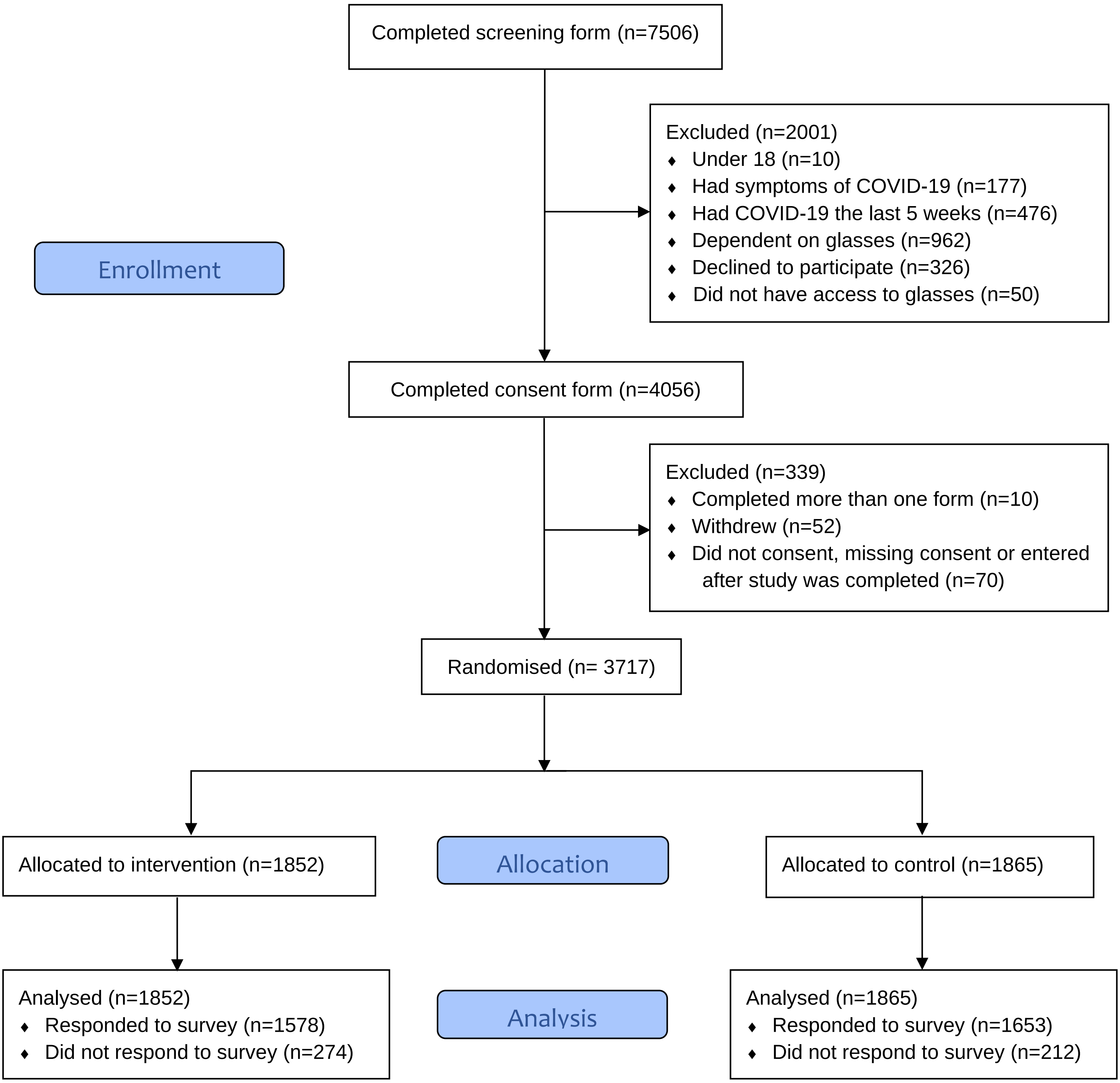
